# Isolated night cough in children: how does it differ from wheeze?

**DOI:** 10.1101/19007500

**Authors:** Maja Jurca, Myrofora Goutaki, Philipp Latzin, Erol A. Gaillard, Ben D. Spycher, Claudia E. Kuehni

## Abstract

**Background:** Children with night cough but no wheeze might have a mild form of asthma (cough variant asthma), sharing risk factors with children who wheeze, and possibly developing wheeze later.

**Methods:** We compared risk factors of children with isolated night cough and children with wheeze in the Leicester Respiratory Cohort study at ages 1, 4, 6, and 9 years. We also compared prognoses of children with isolated night cough, children with wheeze, and asymptomatic children.

**Results:** Among 4,101 children at age 1 year, 2,854 at 4, 2,369 at 6, and 1,688 at 9 years, the prevalence of isolated night cough was 10% at age 1 and 18% in older children, while prevalence of wheeze decreased from 35% at 1 year to 13% at age 9. Although many risk factors were the same for cough and wheeze, day care, reflux, and family history of bronchitis were more strongly associated with cough, and male sex and family history of asthma with wheeze. Over one-third of pre-schoolers with cough continued to cough at school age, but their risk of developing wheeze was similar to that of children asymptomatic at earlier surveys. Wheeze tracked more strongly throughout childhood than cough.

**Conclusions:** Some risk factors for cough and wheeze were shared and some were not; there was little evidence that children with isolated night cough have an increased risk of future wheeze. This suggests that only a fraction of children with isolated night cough might have a variant of asthma, if at all.

## Introduction

Cough is common among children, accounts for many consultations, and can affect quality of life.(1, 2) Most commonly caused by upper or lower respiratory tract infections, cough is usually self-limiting. However, some children cough more frequently, also apart from infections.(3, 4) Many of these children suffer from asthma and experience wheeze and shortness of breath in addition to cough. Others have isolated frequent cough, without wheeze or dyspnoea.(5, 6) In epidemiological studies, these are children who report dry cough at night and cough apart from colds, but no wheeze. It has been postulated, that many of them have cough variant asthma (CVA),(7-10) a phenotype of mild asthma with cough as sole symptom.(11) Patients with CVA have been characterized as having a family or personal history of atopy, eosinophilic inflammation, a high bronchial reactivity, a positive response to bronchodilators, positive exercise provocation tests, cough triggered by exercise, and an increased risk for developing wheeze or typical asthma later.(10, 12-16) The existence of CVA as a disease entity is disputed, though, and several authors have said that isolated cough is a poor marker for wheeze and asthma, and should not be treated as such.(8, 17-20)

The epidemiology of isolated cough in children is not well studied, while risk factors(21-23) and natural history(24, 25) of wheeze have been described extensively. Previous publications on isolated cough were mostly non-systematic reviews,(9, 10, 26) small clinical studies with fewer than 50 patients,(13, 14, 27) or studies that focused on adults.(28-30) Studies that compare children with cough to children with wheeze and asymptomatic children in the same population are scarce.(18, 31, 32) Drawing on the dataset from the Leicester 1998 cohort studies, we compared risk factors of children with isolated cough and those with wheeze, and we compared prognoses of children with isolated cough, children with wheeze, and those with neither symptom.

## Materials and methods

### Study design and population

We analysed data from a large, prospective population-based cohort, the Leicester Respiratory Cohort (LRC).(33) Perinatal and growth data were collected from birth records and health visitor records. The postal questionnaires collected information on cough, wheeze, and environmental exposures from parents when children were aged 1 year (in 1998), and thereafter at the children’s ages of 4 (in 2001), 6 (in 2003), and 9 years (in 2006). This study includes all LRC children who were born between May 1996 and April 1997, and who completed the baseline questionnaire in 1998 (4,101). Among these, 2,854 (70%) returned the questionnaire at age 4 (in 2001), 2,369 (58%) at age 6 (in 2003) and 1,688 (41%) at age 9 (in 2006).

The Leicestershire Health Authority Research Ethics Committee approved the study (approval numbers 1132, 5005, 4867).

### Current wheeze and night cough

We used the questions on current wheeze and dry cough at night, apart from a cough associated with a cold or a chest infection, from the ISAAC key questionnaire.(34) At each survey, we distinguished three mutually exclusive groups of children based on their symptoms during the previous 12 months: children with isolated night cough but no wheeze, children with wheeze with or without cough, and asymptomatic children with neither cough nor wheeze.

### Risk factors

We compiled a list of potential risk factors for cough and wheeze from the literature.(18, 23, 31, 35) These included demographic factors (sex, ethnicity), family (parental) asthma, bronchitis, hay fever, and eczema, exposure to infections (household crowding, day care attendance, older siblings), environmental exposures (cooking with gas, tobacco smoke exposure, pet ownership), socioeconomic factors (maternal education, Townsend deprivation index(36)), and perinatal/early life factors (gestational age, birth weight, maternal age, breastfeeding, reflux in infancy). We also considered clinical data at three surveys—atopic diseases (rhinitis, hay fever, eczema) and ear, nose, and throat (ENT) symptoms (frequent colds, snoring, otitis)—as potential predictors of wheeze and cough at the next survey.

Attendance at day care, reflux, ethnicity, family history, older siblings, and perinatal and early life factors were only assessed at age 1 in 1998.

### Statistical analyses

We calculated the prevalence of isolated cough and wheeze at ages 1, 4, 6, and 9 years, and then investigated risk factors for isolated cough and wheeze at each age using multinomial logistic regression. We calculated univariable relative risk ratios (RRRs) with 95% confidence intervals (CIs) for isolated cough and for wheeze compared to the reference category of asymptomatic children. In adjusted models, we included all risk factors that were associated with either cough or wheeze in the univariable model (p<0.1). We used Wald tests to compare whether RRRs differed between cough and wheeze.

We then investigated the prognoses of children with isolated cough, wheeze, and none of these symptoms by calculating the proportion of children who had the same phenotype at the next survey, and of those who transitioned to another phenotype. We did this for the three different periods from ages 1 to 4, 4 to 6, and 6 to 9 years. We used an overall chi-square test of independence between symptoms at baseline and symptoms at follow-up to assess whether prognosis differed between children with isolated cough, children with wheeze, and children with none of the symptoms. We subsequently performed a subgroup analysis to test whether children with isolated cough had a higher risk of developing wheeze than asymptomatic children using Fisher’s exact test.

Finally, we assessed potential predictors of prognosis, i.e. persistence of cough or incidence of wheeze 2-3 years later, in children who had cough at baseline. We fitted univariable and adjusted models including all variables that were associated in the univariable model (p<0.1). In addition to environmental factors, we also investigated whether clinical data at baseline (atopic diseases and ENT symptoms) helped to predict outcome at the next survey.(37, 38) The layout of our study is shown in **Figure 1**.

**Fig 1.**
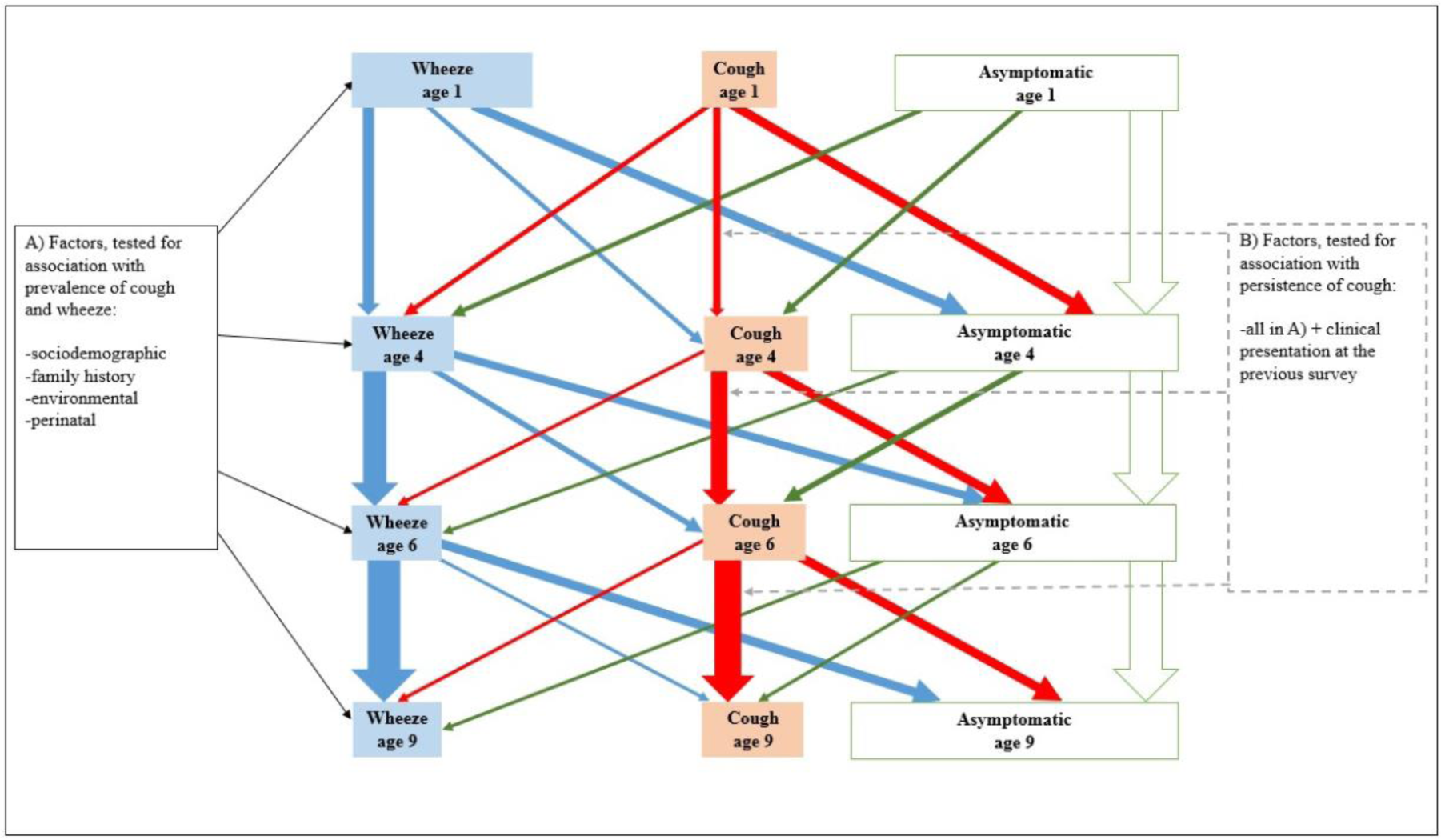
Study design: prevalence of wheeze and isolated cough at ages 1, 4, 6, and 9 years, and factors associated with prevalence and prognosis of these symptoms. The size of rectangular boxes represents the respective number of children with cough, wheeze, or no symptoms, and the width of the arrows semiquantitatively demonstrates the likelihood to stay in the same or change to a different group at subsequent surveys.

We performed sensitivity analyses to test the robustness of our findings. While the main analysis included all children, who participated in the baseline survey at the age of 1 and one or more of the following surveys at 4, 6, or 9 years, the sensitivity analysis included only children who participated in all four surveys (1,318 children, 32% of 4,101). Results from this sensitivity analysis were very similar to the main analysis (available from the authors). We prepared and analysed the data using Stata 14.0 (Stata Corporation LP, Austin, Texas, USA) and created the figures using R version 3.1.1 (Free Software Foundation, Boston, USA).

## Results

The characteristics of the 4,101 participants of the 1998 survey are shown in **Table 1**. Prevalence of isolated night cough was 10% in 1-year-olds and 18% in 4-, 6- and 9-year-olds (**S1 Table**). Prevalence of wheeze declined from 35% in 1-year-olds to 17%, 14%, and 13% respectively in children aged 4, 6, and 9 years.

**Table 1.** Characteristics of the study population at the age of 1 year, in 1998 (N=4,101).

|  | N | % |
| --- | --- | --- |
| **Demographic factors** |  |  |
| Ethnicity: South Asian | 801 | 20 |
| White | 3,300 | 80 |
| Sex: Boys | 2,135 | 52 |
| Girls | 1,966 | 48 |
| **Family history<sup>a</sup>** |  |  |
| Asthma | 1,294 | 32 |
| Bronchitis | 739 | 18 |
| Hay fever | 1,846 | 45 |
| Eczema | 1,339 | 33 |
| **Indoor exposures** |  |  |
| Day care | 1,030 | 25 |
| Older siblings | 2,276 | 58 |
| Cooking with gas | 3,051 | 74 |
| Mother smoking | 895 | 22 |
| Father smoking | 1,041 | 26 |
| Any pets | 1,664 | 41 |
| **Socioeconomic factors** |  |  |
| Low maternal education<sup>b</sup> | 1,956 | 48 |
| Deprived (Townsend score $\geq 1.86$)<sup>c</sup> | 1,303 | 32 |
| **Perinatal and early life data** |  |  |
| Preterm (gestational age <37 weeks) | 281 | 7 |
| Low birth weight (<2500g) | 293 | 7 |
| Young mother (<25 years)<sup>d</sup> | 929 | 23 |
| Child breastfed | 2,432 | 60 |
| Reflux (possetting) in infancy | 3,061 | 75 |
| **Respiratory problems in the past year** |  |  |
| Any night cough | 938 | 23 |
| Any wheeze | 1,420 | 35 |
| Night cough without wheeze | 427 | 10 |
| Cough triggered by |  |  |
| exercise | 408 | 11 |
| aeroallergens | 77 | 2 |
| laughter | 727 | 24 |
| food | 427 | 12 |
| Rhinitis | 1,291 | 32 |
| Eczema<sup>e</sup> | 1,155 | 36 |
| Frequent colds (>6 episodes) | 777 | 19 |
| Snoring | 2,236 | 57 |
| Otitis media | 1,706 | 42 |

<sup>a</sup> Either mother or father with a history of the disease
<sup>b</sup> Age at the end of education of mother is $\leq 16$ years
<sup>c</sup> Townsend Deprivation Index (deprived, the highest 2 quintiles): deprived [1.86, 5.15], more deprived [5.16, 11.07]
<sup>d</sup> When the child was born
<sup>e</sup> Asked only in part of the cohort
Hay fever was not inquired about in 1998 survey.

### Risk factors for prevalent cough and wheeze

Some risk factors were shared between children with isolated night cough and wheeze, others differed, and risk factors also varied with age. Results are shown in **Table 2** for children aged 1 year and **S2-S4 Tables** for children aged 4, 6, and 9 years. The tables report RRRs for children with cough and wheeze compared to asymptomatic children and similarity p-values, which indicate the difference in strength of association between children with wheeze and those with isolated cough. **Figures 2-4** and **S1 Figure** summarize the same results graphically.

**Table 2.** Risk factors for prevalence of isolated night cough and wheeze in 1-year-old children (N=4,101). Association of different factors with cough and wheeze, compared to asymptomatic children, in an unadjusted and adjusted model presented as relative risk ratio estimates with confidence intervals. Cough was defined as night cough without wheeze.

| Risk factors at age 1 | Unadjusted model |  |  | Adjusted model<sup>a</sup> |  |  |
| --- | --- | --- | --- | --- | --- | --- |
|  | Night cough<sup>b</sup> | Wheeze | Similarity p-value<sup>c</sup> | Night cough<sup>b</sup> | Wheeze | Similarity p-value<sup>c</sup> |
|  | RRR (95% CI) | RRR (95% CI) |  | RRR (95% CI) | RRR (95% CI) |  |
| South Asian ethnicity | **1.3 (1.0-1.6)** | **0.6 (0.5-0.8)** | **<0.001** | **1.4 (1.0-2.0)** | **0.8 (0.6-1.0)** | **0.001** |
| Male sex | 0.9 (0.7-1.1) | **1.3 (1.1-1.5)** | **0.003** | 0.9 (0.7-1.2) | **1.3 (1.2-1.6)** | **0.003** |
| Family history of: |  |  |  |  |  |  |
| Asthma | 1.0 (0.8-1.3) | **2.1 (1.8-2.4)** | **<0.001** | 0.9 (0.7-1.2) | **1.7 (1.5-2.0)** | **<0.001** |
| Bronchitis | **1.4 (1.0-1.8)** | **2.0 (1.7-2.3)** | **0.008** | **1.4 (1.0-1.9)** | **1.6 (1.3-2.0)** | 0.380 |
| Hay fever | **1.2 (1.0-1.4)** | **1.5 (1.3-1.7)** | **0.049** | 1.1 (0.9-1.4) | **1.2 (1.1-1.4)** | 0.407 |
| Eczema | 1.1 (0.8-1.3) | **1.3 (1.1-1.5)** | 0.109 | 1.0 (0.8-1.3) | 1.0 (0.8-1.1) | 0.702 |
| Exposure to infections |  |  |  |  |  |  |
| Crowding | 1.1 (0.9-1.4) | **1.2 (1.1-1.4)** | 0.322 | 1.1 (0.8-1.4) | 1.1 (0.9-1.3) | 0.961 |
| Day care | **1.6 (1.3-2.0)** | **1.1 (1.0-1.3)** | **0.002** | **1.9 (1.5-2.4)** | **1.3 (1.1-1.6)** | **0.008** |
| Older siblings | 0.9 (0.7-1.1) | **1.2 (1.0-1.4)** | **0.019** | 0.9 (0.7-1.2) | **1.3 (1.1-1.6)** | **0.008** |
| Indoor exposures |  |  |  |  |  |  |
| Cooking with gas | 1.2 (0.9-1.5) | 1.1 (0.9-1.2) | 0.424 | - | - | - |
| Mother smoking | 1.1 (0.8-1.4) | **1.8 (1.6-2.2)** | **<0.001** | 1.1 (0.8-1.5) | **1.4 (1.2-1.7)** | 0.104 |
| Father smoking | 1.0 (0.8-1.3) | **1.3 (1.1-1.5)** | 0.112 | 1.0 (0.8-1.4) | 1.1 (0.9-1.3) | 0.775 |
| Pets | **0.8 (0.7-1.0)** | **1.1 (1.0-1.3)** | **0.006** | 0.9 (0.7-1.2) | 1.1 (0.9-1.2) | 0.273 |
| Socioeconomic factors |  |  |  |  |  |  |
| Low maternal education<sup>d</sup> | 1.0 (0.8-1.3) | **1.3 (1.1-1.4)** | **0.056** | 1.1 (0.9-1.4) | 1.0 (0.9-1.2) | 0.613 |
| Deprivation (Townsend) | **1.4 (1.1-1.7)** | **1.4 (1.2-1.6)** | 0.908 | **1.4 (1.0-1.8)** | **1.5 (1.3-1.8)** | 0.546 |
| Perinatal and early life |  |  |  |  |  |  |
| Preterm (GA<37 weeks) | 0.8 (0.5-1.3) | 1.2 (0.9-1.6) | **0.096** | - | - | - |
| Low birthweight(<2500g) | 0.8 (0.5-1.3) | 1.1 (0.9-1.5) | 0.187 | - | - | - |
| Young mother (<25 yrs)<sup>e</sup> | 1.2 (0.9-1.5) | **1.5 (1.3-1.8)** | **0.048** | 1.1 (0.8-1.5) | **1.4 (1.2-1.7)** | **0.091** |
| Breastfeeding | 1.0 (0.8-1.2) | **0.7 (0.6-0.8)** | **0.021** | 0.9 (0.7-1.2) | 0.9 (0.7-1.0) | 0.485 |
| Reflux in infancy | **1.7 (1.3-2.2)** | **1.4 (1.2-1.7)** | 0.193 | **1.7 (1.3-2.3)** | **1.4 (1.2-1.7)** | 0.163 |

RRR, relative risk ratio; CI, confidence interval; GA, gestational age
<sup>a</sup> Adjusted model includes all covariates with p-values <0.10 for either cough or wheeze in univariable models; baseline for multinomial regression: asymptomatic children
<sup>b</sup> Defined as night cough without wheeze (ISAAC questions)
<sup>c</sup> p-value from test for difference between associations of risk factors with cough and those with wheeze (Wald test)
<sup>d</sup> End of education of mother at age <16 years
<sup>e</sup> When the child was born

**Fig 2.**
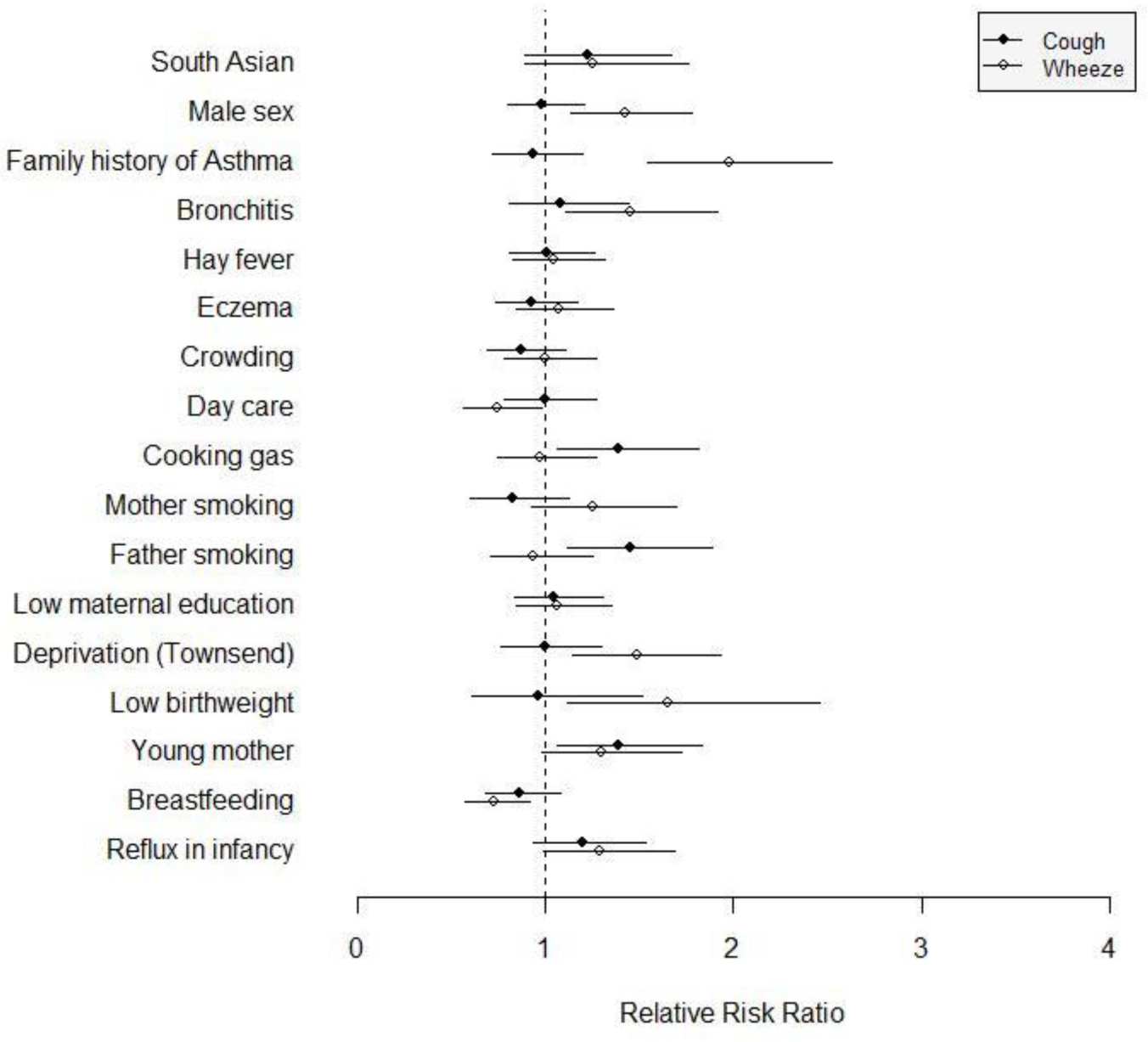
Risk factors for prevalent isolated night cough and wheeze at age 4 year (N=2,854). Association of different factors with cough and with wheeze, compared to asymptomatic children, in a fully adjusted model (adjusted for all covariates with p-values <0.10 for either cough or wheeze in univariable models), presented as relative risk ratio estimates with confidence intervals. Cough was defined as night cough without wheeze.

**Fig 3.**
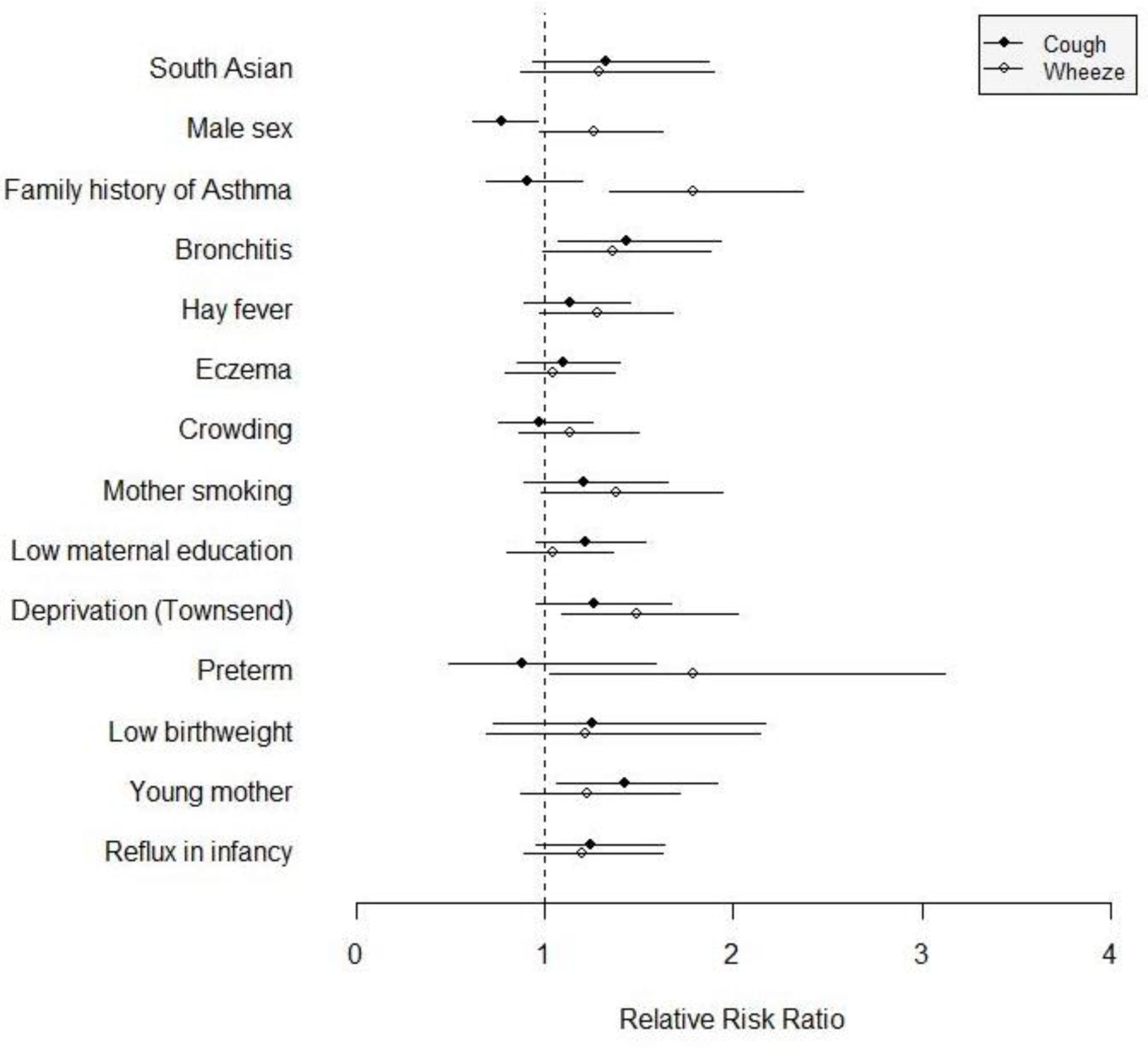
Risk factors for prevalent isolated night cough and wheeze at age 6 year (N=2,369). Association of different factors with cough and with wheeze, compared to asymptomatic children, in a fully adjusted model (adjusted for all covariates with p-values <0.10 for either cough or wheeze in univariable models), presented as relative risk ratio estimates with confidence intervals. Cough was defined as night cough without wheeze.

The risk factors that were shared for isolated cough or wheeze were family history of bronchitis, low socioeconomic status, exposure to smoking, and reflux (posseting or vomiting in infancy). Factors that were associated mainly with cough were South Asian ethnicity, day care attendance (age 1), paternal smoking, and use of gas for cooking (age 4). Several factors were more important for wheeze. Boys had a higher risk for wheeze at all ages, but a lower risk for cough at ages 6 and 9. A family history of asthma or hay fever was associated with wheeze only. Maternal smoking and presence of older siblings were more strongly associated with wheeze at age 1. Low birthweight, preterm birth, and lack of breastfeeding were associated mainly with wheeze. Similarity p-values suggested that associations differed significantly between wheeze and cough for sex and family history of asthma at all ages; ethnicity, day care attendance, and older siblings at age 1; and cooking with gas, exposure to smoking, low socioeconomic status, and preterm birth at age 4 (**Table 2, S2-S4 Tables**).

### Prognosis of isolated night cough and wheeze

Prognosis differed between children with wheeze, those with isolated night cough, and those who had no such symptoms - for all intervals studied from ages 1 to 4, 4 to 6, and 6 to 9 years (overall p-values <0.001, **Figure 5, S5 Table**). The proportion of children with isolated cough, whose cough persisted, increased from 32% (99/305) between age 1 and 4, to 42% (160/381) from age 4 to 6, and 39% (97/249) from age 6 to 9 years. Wheeze was more persistent than cough. Among children with wheeze at age 1, 31% (283/921) wheezed at age 4 years. Among 4-year-olds with wheeze, 48% (151/315) wheezed again at age 6, and among 6-year-olds with wheeze, 59% (98/165) reported wheeze at age 9.

**Fig 4.**
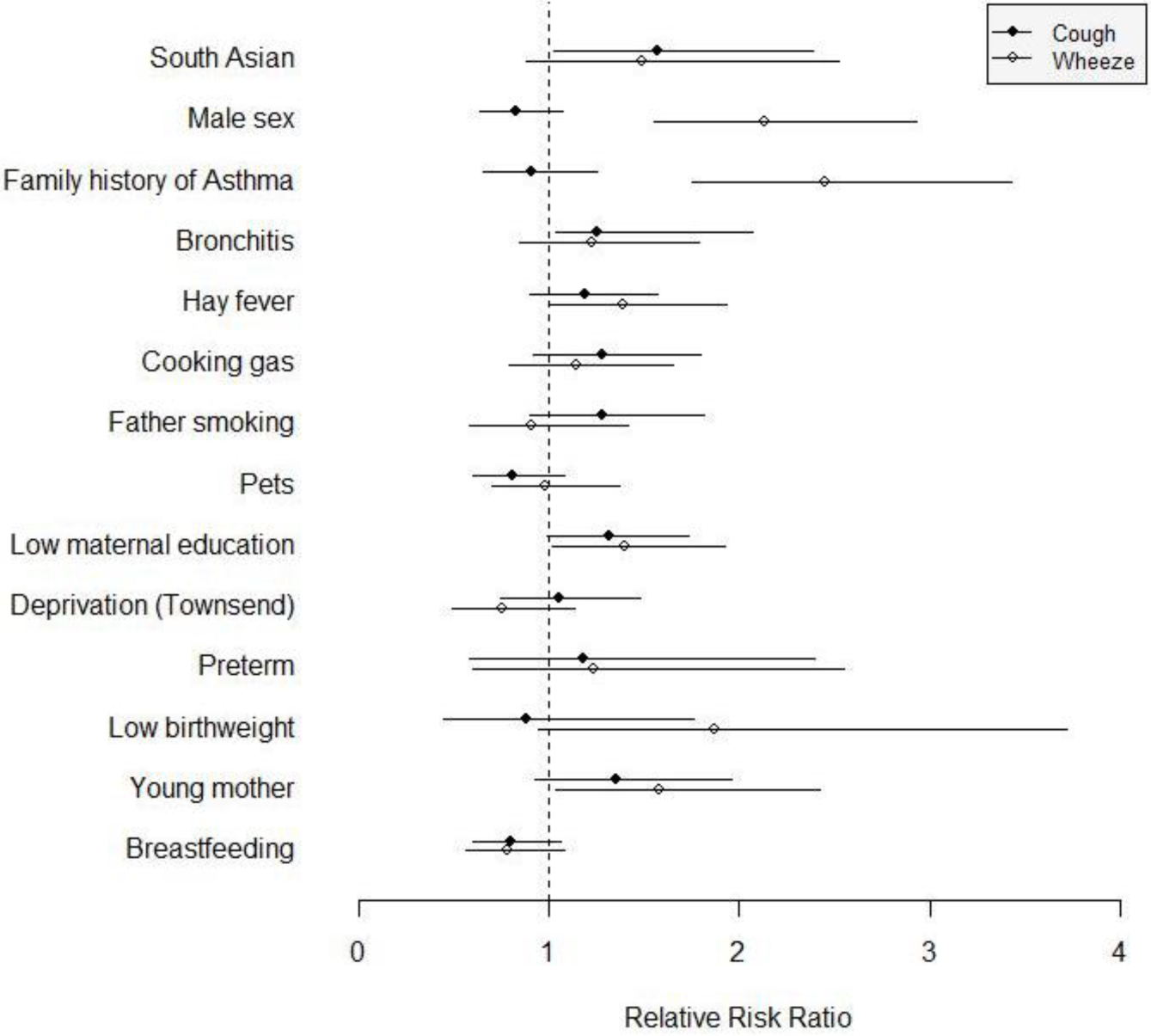
Risk factors for prevalent isolated night cough and wheeze at age 9 year (N=1,688). Association of different factors with cough and with wheeze, compared to asymptomatic children, in a fully adjusted model (adjusted for all covariates with p-values <0.10 for either cough or wheeze in univariable models), presented as relative risk ratio estimates with confidence intervals. Cough was defined as night cough without wheeze.

**Fig 5.**
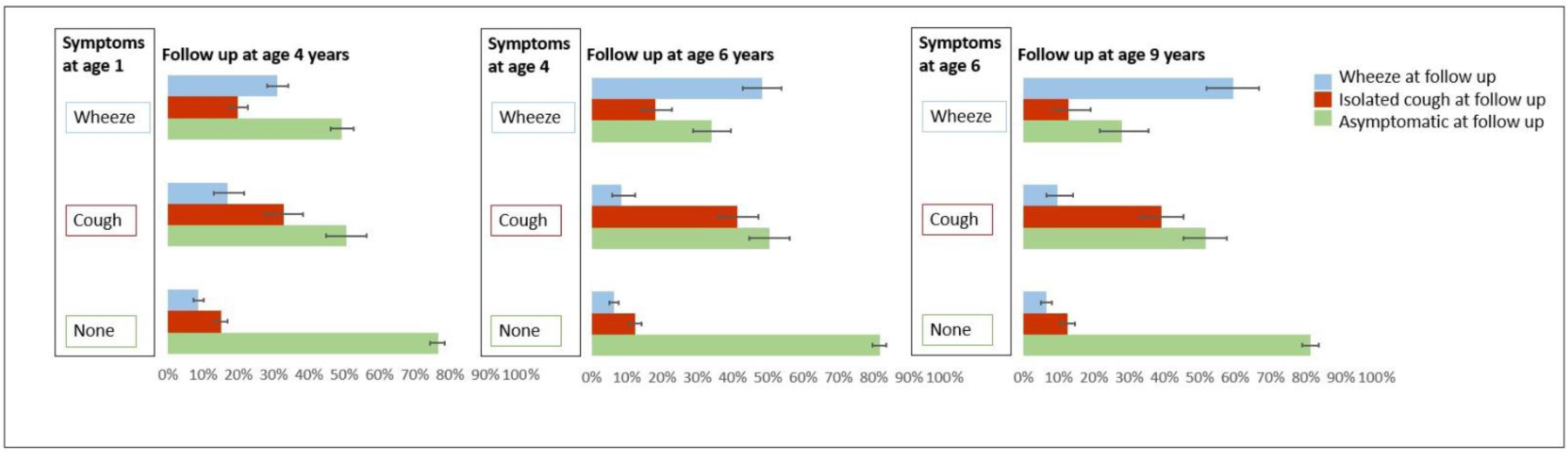
Prognosis and tracking of respiratory symptoms in children. Proportion of children with wheeze, isolated night cough, or none of the symptoms, who have wheeze, cough, or are asymptomatic 2-3 years later. This figure shows prognosis for 3 different development periods in childhood (1 to 4 years, 4 to 6 years, 6 to 9 years).

The risk of developing new (incident) wheeze at the next survey was not higher in children with isolated cough than in asymptomatic children, with the possible exception of infants, among whom 17% with isolated night cough reported wheeze 3 years later compared to 9% in asymptomatic children (p<0.001). In children aged 4 or 6 years, we found no evidence that the risk of wheeze at the next survey differed in those with isolated cough and those with neither cough nor wheeze.

### Predictors of future symptoms in children with cough

Factors associated with prognosis of isolated night cough are shown in **Table 3**. Some factors were associated with both persistent cough and incident wheeze, usually with stronger effect sizes for later wheeze such as cough triggered by laughter (4 to 6) or by aeroallergens (6 to 9) and hay fever (6 to 9). Persistence of cough was not strongly associated with any of the tested environmental or perinatal factors from ages 1 to 4, 4 to 6 or 6 to 9 years (**Table 3**). Clinical factors helped predict persistent cough in older children: from age 4 to 6 years, rhinitis, cough triggered by laughter/crying, and snoring predicted persistence of isolated night cough; from 6 to 9 years, children with frequent infections (frequent colds, otitis) or allergies (hay fever, cough triggered by aeroallergens) were more likely to have persistent cough 3 years later.

**Table 3.** Predictors of future symptoms in children with isolated night cough at baseline for different age intervals (from age 1 to 4 years, from age 4 to 6 years, and from age 6 to 9). Example for the youngest age group: In children with isolated night cough at age 1, what are the factors associated with persistence of isolated cough vs. incidence of wheeze at age 4 years? The presented associations are univariable.

| Risk factors at baseline | From age 1 to 4 years (N=305) |  |  | From age 4 to 6 years (N=381) |  |  | From age 6 to 9 years (N=249) |  |  |
| --- | --- | --- | --- | --- | --- | --- | --- | --- | --- |
|  | Night cough<sup>a</sup> | Wheeze | Similarity<br>p-value<sup>b</sup> | Night cough<sup>a</sup> | Wheeze | Similarity<br>p-value<sup>b</sup> | Night cough<sup>a</sup> | Wheeze | Similarity<br>p-value<sup>b</sup> |
|  | RRR (95% CI) | RRR (95% CI) |  | RRR (95% CI) | RRR (95% CI) |  | RRR (95% CI) | RRR (95% CI) |  |
| South Asian ethnicity | 1.0 (0.5-1.8) | 1.3 (0.6-2.7) | 0.412 | 1.2 (0.7-2.2) | 1.8 (0.7-4.5) | 0.380 | 1.1 (0.5-2.2) | 0.7 (0.2-2.5) | 0.486 |
| Male sex | 0.8 (0.5-1.3) | 1.6 (0.9-3.1) | **0.037** | 0.7 (0.5-1.1) | 1.7 (0.8-3.7) | **0.029** | 0.8 (0.5-1.4) | 2.2 (0.9-5.6) | **0.036** |
| *Family history of* |  |  |  |  |  |  |  |  |  |
| Asthma | 1.2 (0.7-2.3) | **3.0 (1.5-6.0)** | **0.018** | 1.3 (0.8-2.0) | **2.3 (1.1-5.0)** | 0.127 | 1.0 (0.5-1.8) | **3.4 (1.4-8.2)** | **0.009** |
| Bronchitis | 0.7 (0.4-1.4) | 1.4 (0.7-3.0) | 0.112 | 1.1 (0.6-1.9) | 1.1 (0.4-2.8) | 0.988 | 1.5 (0.7-2.9) | 1.8 (0.6-5.0) | 0.724 |
| Hay fever | 0.9 (0.6-1.5) | 1.3 (0.7-2.5) | 0.288 | 1.3 (0.8-1.9) | 1.8 (0.9-3.9) | 0.350 | **1.7 (1.0-2.9)** | 1.6 (0.7-3.8) | 0.895 |
| Eczema | 1.2 (0.7-2.0) | 1.0 (0.5-1.9) | 0.598 | 1.4 (0.9-2.2) | 1.8 (0.8-4.0) | 0.472 | 0.6 (0.3-1.0) | 0.8 (0.3-1.9) | 0.538 |
| *Exposure to infections* |  |  |  |  |  |  |  |  |  |
| Crowding | 1.0 (0.6-1.8) | 1.4 (0.7-2.7) | 0.425 | 0.9 (0.5-1.4) | 1.5 (0.7-3.4) | 0.179 | 1.1 (0.6-1.8) | 1.1 (0.4-2.7) | 0.953 |
| Day care at age 1 | 1.2 (0.6-2.3) | 0.3 (0.1-0.7) | **0.003** | - | - | - | - | - | - |
| Older siblings | 0.8 (0.5-1.4) | 0.6 (0.3-1.1) | 0.315 | 1.0 (0.6-1.5) | 0.7 (0.3-1.6) | 0.530 | 1.2 (0.7-2.1) | 1.5 (0.6-3.7) | 0.654 |
| *Air pollution* |  |  |  |  |  |  |  |  |  |
| Cooking with gas | 1.1 (0.6-1.9) | 0.7 (0.4-1.5) | 0.352 | 1.3 (0.7-2.2) | 1.4 (0.5-3.8) | 0.883 | 1.3 (0.7-2.4) | 3.1 (0.9-11.0) | 0.207 |
| Mother smoking | 1.3 (0.7-2.7) | **2.2 (1.0-4.8)** | 0.214 | 1.3 (0.7-2.4) | 0.7 (0.2-2.4) | 0.289 | 1.0 (0.5-2.0) | 1.0 (0.3-3.1) | 0.984 |
| Father smoking | 1.2 (0.7-2.2) | 1.1 (0.5-2.3) | 0.788 | 1.0 (0.6-1.6) | 0.7 (0.3-1.9) | 0.529 | 1.0 (0.5-2.0) | 0.9 (0.3-2.7) | 0.814 |
| *Allergens* |  |  |  |  |  |  |  |  |  |
| Pets | 1.0 (0.6-1.7) | 0.6 (0.3-1.2) | 0.171 | 1.0 (0.7-1.5) | 0.6 (0.3-1.3) | 0.188 | 0.9 (0.6-1.6) | **2.5 (1.0-6.0)** | **0.043** |
| *Socioeconomic factors* |  |  |  |  |  |  |  |  |  |
| Low maternal education<sup>c</sup> | 1.0 (0.6-1.6) | 1.1 (0.6-2.0) | 0.802 | 1.0 (0.6-1.5) | 0.6 (0.3-1.3) | 0.233 | **1.6 (1.0-2.8)** | 2.2 (0.9-5.2) | 0.552 |
| Deprivation(Townsend) | 0.7 (0.4-1.3) | 1.7 (0.9-3.2) | **0.019** | 1.4 (0.9-2.3) | 2.1 (0.9-4.6) | 0.338 | 1.1 (0.6-1.9) | 0.4 (0.1-1.3) | **0.095** |
| *Perinatal and early life* |  |  |  |  |  |  |  |  |  |
| Preterm (GA<37 weeks) | 0.3 (0.03-2.2) | 1.5 (0.4-6.4) | 0.125 | 1.0 (0.4-2.5) | 2.5 (0.7-8.6) | 0.130 | - | - | - |
| Low birthweight(<2500g) | 3.3 (0.8-13.5) | 2.0 (0.3-12.6) | 0.566 | 1.0 (0.4-2.4) | 1.7 (0.4-6.3) | 0.440 | - | - | - |
| Young mother (<25 yrs) | 1.2 (0.6-2.2) | 1.3 (0.6-2.9) | 0.753 | 1.3 (0.8-2.3) | 1.3 (0.5-3.3) | 0.922 | 1.2 (0.6-2.5) | 1.6 (0.5-4.8) | 0.616 |
| Breastfeeding | 0.8 (0.5-1.3) | 0.7 (0.3-1.3) | 0.694 | 1.1 (0.7-1.7) | 0.6 (0.3-1.4) | 0.191 | 0.5 (0.3-0.9) | 0.4 (0.2-1.1) | 0.683 |
| Reflux in infancy | 1.4 (0.7-2.7) | 1.5 (0.6-3.4) | 0.920 | 0.9 (0.6-1.5) | 1.9 (0.6-5.7) | 0.206 | 0.9 (0.5-1.8) | 0.6 (0.2-1.6) | 0.370 |
| *Clinical factors* |  |  |  |  |  |  |  |  |  |
| Rhinitis | 1.2 (0.7-2.0) | 1.0 (0.5-2.0) | 0.640 | **1.6 (1.0-2.5)** | 1.9 (0.9-4.1) | 0.655 | 1.3 (0.8-2.3) | **3.5 (1.4-8.7)** | **0.042** |
| Hay fever<sup>d</sup> | - | - | - | 1.1 (0.5-2.0) | 1.9 (0.7-2.3) | 0.270 | **4.3 (1.6-11.4)** | **5.3 (1.5-19.1)** | 0.722 |
| Current eczema | 1.0 (0.5-1.9) | 1.5 (0.7-3.1) | 0.335 | 1.3 (0.6-1.9) | 2.1 (0.8-5.3) | 0.175 | 1.6 (0.9-3.0) | **3.4 (1.4-8.4)** | 0.110 |
| Cough triggered by |  |  |  |  |  |  |  |  |  |
| exercise | 1.1 (0.5-2.7) | **3.8 (1.7-8.8)** | **0.010** | 1.3 (0.8-2.1) | 1.9 (0.8-4.2) | 0.409 | 1.4 (0.7-2.5) | 4.1 (0.5-10.9) | **0.034** |
| aeroallergens | 0.3 (0.03-2.7) | 1.9 (0.4-8.0) | 0.123 | 1.4 (0.7-2.8) | **3.0 (1.1-8.0)** | 0.113 | **5.2 (2.0-13.6)** | **8.3 (2.5-27.6)** | 0.370 |
| laughter | **2.3 (1.2-4.4)** | 1.2 (0.5-2.8) | 0.138 | **1.7 (1.0-2.8)** | **4.1 (1.5-11)** | **0.071** | 1.7 (0.9-3.3) | 1.0 (0.3-3.0) | 0.343 |
| food | 1.3 (0.7-2.6) | 0.7 (0.2-1.8) | 0.187 | 1.0 (0.5-1.9) | 1.0 (0.3-3.6) | 0.953 | 1.9 (0.8-4.6) | 0.5 (0.1-4.5) | 0.237 |
| Frequent colds<sup>e</sup> | 0.8 (0.4-1.4) | 0.9 (0.4-2.0) | 0.640 | 1.1 (0.5-2.3) | 1.0 (0.3-3.8) | 0.906 | **4.8 (1.3-17.8)** | 1.8 (0.2-18.0) | 0.366 |
| Snoring | 0.8 (0.5-1.4) | **2.1 (1.0-4.4)** | **0.020** | **1.9 (1.1-3.0)** | 2.1 (0.9-5.2) | 0.777 | 1.3 (0.7-2.3) | **3.4 (1.1-10.6)** | 0.105 |
| Otitis at least once | 1.1 (0.7-1.9) | 0.9 (0.5-1.7) | 0.437 | 1.0 (0.7-1.6) | 1.4 (0.7-3.0) | 0.408 | **2.1 (1.2-3.7)** | 1.6 (0.6-3.9) | 0.546 |

RRR, relative risk ratio; CI, confidence interval; GA, gestational age
<sup>a</sup> Cough defined as night cough and no wheeze
<sup>b</sup> p-value from test for difference between associations of risk factors with cough and those with wheeze (Wald test)
<sup>c</sup> End of education of mother at age <16 years
<sup>d</sup> Not inquired about in 1998 survey
<sup>e</sup> >6 episodes of colds in the past year

Some factors were associated with progression from isolated cough to wheeze, particularly in older children. Children with isolated night cough who had a family history of asthma had an increased risk of later wheeze in all age groups. Children whose cough was triggered by aeroallergens or by laughter/crying at age 4 years had an increased risk of wheeze at age 6.

Children who had also rhinitis, hay fever, eczema, cough triggered by aeroallergens, snoring or exposure to pets at age 6 years had an increased risk of incident wheeze at age 9 years (**Table 3**).

## Discussion

In this large population-based study, which compared risk factors and prognosis of isolated night cough with wheeze in different age groups from infancy to school age, some risk factors were shared for cough and wheeze, while others differed. Persistence of symptoms was stronger for wheeze than for cough and was higher in older children. Overall, we found no clear evidence that isolated night cough progressed to wheeze.

Risk factors for cough have been little studied, mainly in clinical settings,(1, 39, 40) and rarely in the general population.(18, 31) A cross-sectional Danish study of 2,978 5-year-olds reported male sex, low gestational age, maternal asthma, and housing conditions as risk factors for wheeze, but not for isolated cough; it proposed that the aetiology differs for the two phenotypes.(31) The Tucson Children’s Respiratory Study of 987 6-year-olds, identified parental history of bronchitis and passive smoking as risk factors for cough without colds.(18) Passive smoking and day care attendance have been reported as risk factors for isolated cough by other studies.(41-43) A Finish study reported that 7 to 12-year-olds with isolated night cough had an intermediate prevalence of parental asthma and allergies compared to children with wheeze and asymptomatic children, and suggested that this supports the existence of CVA.(44) In contrast, a cross-sectional Australian study of 1,165 schoolchildren found no differences between children with persistent isolated cough and asymptomatic children with respect to family history of asthma or allergy, parental smoking, and atopic status, and concluded that children with isolated cough are unlikely to have asthma.(17) In our study we found risk factors that were similar for cough and wheeze, like family history of bronchitis and reflux, and others that differed (including ethnicity, sex, family history of asthma, and day care attendance).

Prognosis of isolated cough in children also has received little attention in previous studies. We found that both cough and wheeze tracked more strongly with increasing age. As depicted by the width of the arrows, **Figure 1** shows how wheeze tracked more than cough. But we found no evidence that the risk of developing wheeze was higher in children with isolated cough than it was in asymptomatic children. In the first LRC (482 children born 1985-90, no overlap with our study population), 37% of 3-year-olds with cough without colds continued to cough at age 6 years, and 7% developed wheeze.(45) This is similar to what we found from age 4 to 6 years in the second LRC: 42% continued coughing and 8% developed wheeze.

Some studies have investigated whether children with night cough progress to wheeze or asthma, but did not assess persistence of cough.(46-48) Among the 3,252 children of the PIAMA birth cohort, less than 10% of children with isolated night cough at age 2-7 years developed asthma at age 8, and this was comparable to asymptomatic children.(46) The Tucson Children’s Respiratory Cohort found that cough without colds at age 2 persisted in 40% of children by age 6, and in 35% from age 6-11. Risk factors for cough persistence were parental smoking from age 2-6 and atopy from age 6-11.(18) We found that schoolchildren with isolated cough have an increased risk for future wheeze if they also suffer from rhinitis, hay fever, or cough triggered by known asthma triggers. Overall, though, children with isolated cough did not have a substantially higher risk of developing wheeze than asymptomatic peers.

This is the first study to compare risk factors and prognosis among children with isolated cough and children with wheeze in a large childhood cohort from the general population. It employed consistent methodology with standardized questions to assess symptoms and environmental factors in four surveys. It is also the first study to have investigated factors that predict persistence of cough and incidence of wheeze. Among its limitations is its dependence on parental reports, which may be prone to reporting bias.(49) This reflects however a reality in primary care: the physician, too, must rely upon parental reporting of symptoms.

What do children with isolated night cough have in common with children who wheeze? The possibility that CVA underlies isolated cough was a principal motivator of this study. While CVA may be characterized with information on atopy, response to asthma medication, bronchial reactivity, and later wheeze, our study could assess only parent-reported symptoms and risk of later wheeze. We did not have information on bronchial hyper-responsiveness and response to bronchodilators.

Our overall finding that risk factors for cough and wheeze in children only partially overlap suggests that children with isolated night cough do not have an increased risk of future wheeze. Only a fraction of children with isolated night cough, if any, might have a variant of asthma.

## Data Availability

All relevant data are within the manuscript and its Supporting Information files.

## Acknowledgements

We thank the cohort participants and the parents of the Leicester Respiratory Cohorts for completing the questionnaires. We thank Garyfallos Konstantinoudis (ISPM, University of Bern, Switzerland) for his contribution to the preparation of the figures. We thank Christopher Ritter (ISPM, University of Bern, Switzerland) for his editorial assistance.

## Author contributions

Conceptualization: C.E.K., M.J. Data Curation: C.E.K., E.A.G. Formal analysis: M.J., B.D.S. Funding Acquisition: C.E.K., B.D.S., P.L., E.A.G. Investigation: C.E.K., M.J., B.D.S., P.L., M.G. Methodology: C.E.K., M.J., B.D.S. Project Administration: C.E.K., M.J. Supervision: C.E.K. Visualization: M.J. Writing – Original Draft Preparation: M.J., C.E.K. Writing – Review & Editing: C.E.K., M.J., M.G., B.D.S., P.L., E.A.G.

## Supporting information captions

**S1 Table. Prevalence of respiratory symptoms at ages 1, 4, 6, and 9 years**.

**S2 Table. Risk factors for prevalent isolated night cough and wheeze in 4-year-old children (N=2**,**854)**. Association of different factors with cough and wheeze, compared to asymptomatic children, in an unadjusted and adjusted model presented as relative risk ratio estimates with confidence intervals. Cough was defined as isolated night cough (without wheeze).

**S3 Table. Risk factors for prevalent isolated night cough and wheeze in 6-year-old children (N=2**,**369)**. Association of different factors with cough and wheeze, compared to asymptomatic children, in unadjusted and adjusted models presented as relative risk ratio estimates with confidence intervals. Cough was defined as isolated night cough (without wheeze).

**S4 Table. Risk factors for prevalent isolated night cough and wheeze in 9-year-old children (N=1**,**688)**. Association of different factors with cough and wheeze, compared to asymptomatic children, in unadjusted and adjusted models presented as relative risk ratio estimates with confidence intervals. Cough was defined as isolated night cough (without wheeze).

**S1 Fig. Risk factors for prevalent isolated night** cough **and wheeze at age 1 year (N=4**,**101)**. Association of different factors with cough and with wheeze, compared to asymptomatic children, in a fully adjusted model (adjusted for all covariates with p-values <0.10 for either cough or wheeze in univariable models), presented as relative risk ratio estimates with confidence intervals. Cough was defined as night cough without wheeze.

**S5 Table. Prognosis of isolated night cough and wheeze from age 1-4 years, 4-6 years, 6-9 years**. Prognosis shown for all children who were aged 1 in 1998, and who replied to the questionnaires in the respective surveys. Cough was defined as night cough without wheeze.

### Abbreviations

CVA: cough variant asthma
LRC: Leicester Respiratory Cohort
UK: United Kingdom
ISAAC: International Study of Asthma and Allergies in Childhood
ENT: ear, nose and throat
RRR: relative risk ratio
CI: confidence interval
PIAMA: Prevention and Incidence of Asthma and Mite Allergy

